# Machine Learning Prediction of Pharmacogenetic Test Uptake Among Opioid-Prescribed Patients Using Electronic Health Records: A Retrospective Cohort Study

**DOI:** 10.1101/2025.09.26.25336591

**Authors:** Mohammad Yaseliani, Je-Won Hong, Jiang Bian, Larisa Cavallari, Julio Duarte, Danielle Nelson, Wei-Hsuan Lo-Ciganic, Khoa Anh Nguyen, Md Mahmudul Hasan

## Abstract

**Background:** Opioids are a widely prescribed class of medication for pain management. However, they have variable efficacy and adverse effects among patients, due to complex interplay between biological and clinical factors. Pharmacogenetic (PGx) testing can be utilized to match patients’ genetic profiles to individualize opioid therapy, improving pain relief and reducing the risk of adverse effects. Despite its potential, PGx uptake—utilization of PGx testing—remains low due to a range of barriers at the patient, health care provider, infrastructure, and financial levels. Since testing typically involves a shared decision between the provider and patient, predicting likelihood of patient undergoing PGx testing and understanding the factors influencing that decision can help optimize resource use and improve outcomes in pain management.

**Objective:** To develop machine learning (ML) models, identifying patients’ likelihood of PGx uptake based on their demographics, clinical variables, medication use, and social determinants of health (SDoH).

**Methods:** We utilized electronic health records (EHR) data from a single center healthcare system to identify patients prescribed opioids. We extracted patients’ demographics, clinical variables, medication use, and SDoH, and developed and validated ML models, including neural networks (NN), logistic regression (LR), random forests (RF), gradient boosting (XGB), naïve bayes (NB), and support vector machines (SVM) for PGx uptake prediction based on procedure codes. We performed 5-fold cross validation (CV) and created an ensemble probability-based classifier using the best-performing ML models for PGx uptake prediction. Various performance metrics, uptake stratification analysis, and feature importance analysis were employed to evaluate the performance of the models.

**Results:** The ensemble model using XGB and SVM-RBF classifiers had the highest C-statistics at 79.61%, followed by XGB (78.94%), and NN (78.05%). While XGB was the best-performing model, the ensemble model achieved a high accuracy (67.38%), recall (76.50%), specificity (67.25%), and negative predictive value (99.49%). The uptake stratification analysis using the ensemble model indicated that it can effectively distinguish across uptake probability deciles, where those in the higher strata are more likely to undergo PGx in real-world (6.59% in the highest decile compared to 0.12% in the lowest). Furthermore, SHAP value analysis using the XGB model indicated age, hypertension, and household income as the most influential factors for PGx uptake prediction.

**Conclusions:** The proposed ensemble model demonstrated a high performance in PGx uptake prediction among patients using opioids for pain. This model can be utilized as a decision support tool, assisting clinicians in identifying patients’ likelihood of PGx uptake and guiding appropriate decision-making.

## 1. Introduction

Although effective for many, opioid prescribing presents a complex therapeutic challenge due to variable efficacy and the risk of adverse effects often resulting in a trial-and-error approach to opioid prescribing. These variations in opioid response often arise from interindividual differences in pharmacokinetics and pharmacodynamics, influenced in part by genetic variations [1]. Over recent decades, pharmacogenetic testing (PGx) has emerged as a promising strategy to tailor opioid therapy to an individual genetic profile, with the goal of enhancing pain relief, minimizing adverse effects, and improving overall patient outcomes. Several opioids, including codeine, tramadol, hydrocodone, oxycodone are metabolized—processed by the body–to varying extents by CYP2D6, an enzyme involved in the metabolism of many drugs. CYP2D6 is a highly polymorphic gene with over 130 identified variants that can result in varying enzyme activity, categorized into four phenotypes: poor metabolizer, intermediate metabolizer, normal metabolizer, and ultrarapid metabolizer. These phenotypes can influence the efficacy and side effects of drugs metabolized by CYP2D6 [2]. A pragmatic trial demonstrated improved composite pain intensity outcomes with CYP2D6-guided pain management, highlighting the potential benefits of personalized therapy based on pharmacogenetic testing [2]. Based on current evidence, the Clinical Pharmacogenetics Implementation Consortium (CPIC) provides guidelines on using CYP2D6 genotype result for prescribing tramadol, hydrocodone and codeine [2].

Despite increasing evidence supporting the benefits of PGx in improving patient outcome, its routine implementation faces significant challenges at multiple levels, including patient, healthcare provider, infrastructure, and financial barriers. A major obstacle is the knowledge gap among healthcare professionals. Many providers lack sufficient training in pharmacogenetics, hindering their ability to effectively integrate genetic testing into their prescribing practice [3]. This educational deficit is further complicated by the absence of standardized clinical workflows to incorporate pharmacogenetic data seamlessly [4, 5]. Additionally, regional- and patient-level social determinants of health (SDoH), such as socioeconomic status, insurance coverage, and access to healthcare, can limit patient access to PGx testing and personalized care [6]. As a result, PGx remains underutilized in opioid prescribing. Predicting a likelihood of patient undergoing PGx testing and identifying the underlying factors influencing that decision is crucial for enhancing the clinical utility and cost-effectiveness of PGx. Since testing often involves a shared decision between the provider and patient following a discussion of potential benefits and risks, understanding these dynamics can help healthcare providers develop targeted strategies to encourage appropriate use of PGx testing. This, in turn, can lead to improved patient outcomes in pain management [7].

Predicting PGx uptake using electronic health records (EHRs) presents significant challenges because of the complexity and diversity of clinical data, including, laboratory results, genetic information, and patient-reported outcomes. Additionally, demographics and SDoH—such as race, gender, socioeconomic status, environmental exposures, access to healthcare, and lifestyle choices—significantly impact PGx uptake [8]. These diverse data sources provide valuable insights but also pose substantial challenges because their relationships with PGx uptake are often non-linear and complex [9]. Traditional regression models assume linear relationships, which limits their predictive accuracy in this context. This limitation can lead to inaccurate predictions, as conventional methods fail to capture the intricate nonlinear relationships present in complex data. Machine learning (ML) techniques such as neural networks (NN), random forests (RF), and logistic regression (LR) have been increasingly employed to enhance predictive accuracy by identifying and understanding nonlinear relationships within high-dimensional data [10]. While ML has gained significant attention in medicine, particularly in pharmacogenomics, its application has primarily focused on analyzing genetic or proteomic data to identify patterns associated with drug response [11]. This narrow focus overlooks the potential value of integrating a broader range of relevant data types to better facilitate PGx uptake. By leveraging ML algorithms to process extensive and complex data, we can more accurately identify patients with the highest likelihood of undergoing PGx, enabling a more personalized and effective approach to opioid therapy.

In this study, we aimed to develop and validate ML algorithms to identify opioid-prescribed patients who are most likely to undergo PGx test. By analyzing clinical data, demographics, medication use, and SDoH, our models seek to identify factors influencing the uptake of PGx in opioid therapy. We hypothesize that ML-driven predictions can significantly enhance the targeted use of PGx in patients receiving opioids, leading to improved pain management and optimized therapeutic outcomes. Ultimately, our models may facilitate better utilization of PGx testing for opioid use in pain management. This paper outlines our methodological approach, findings, and potential implications for clinical practice in the field of pain management.

## 2. Method

The overall ML pipeline for PGx uptake prediction is shown in Fig. 1. The pipeline involves four steps, including study design, data pre-processing and feature selection, model development, and comprehensive evaluation. Each step involves several tasks that are critical to building and evaluating the final predictive model. The details of each step are described in this section.

**Fig. 1.**
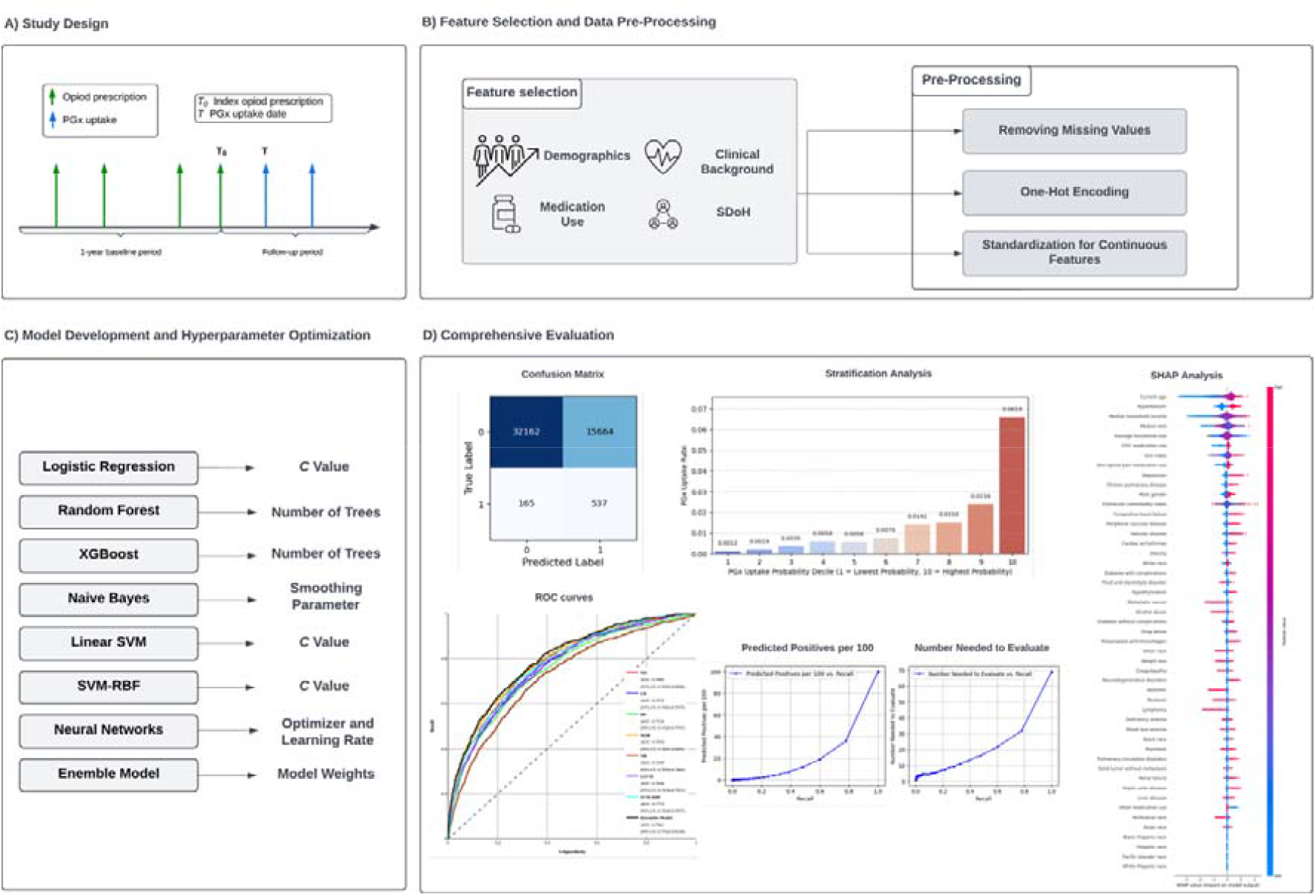
Overall ML pipeline for PGx uptake prediction.

### 2.1. Data source and study design

We utilized real world electronic health record (EHR) data available from the University of Florida Health Integrated Data Repository (UF Health IDR) to develop the models. Patients’ age range was 18 to 89 years, and we did not exclude any patients based on age. Patients were included if they were between 18 and 89 years had an opioid prescription order for non-cancer treatment from 2011 to 2020. Patients were stratified into an intervention group (patient with a minimum of one PGx order) and control group (those with no history of PGx order). We used current procedural terminology (CPT) codes to determine patients’ PGx test. To create the cohort for ML model development, we defined the index dates separately for intervention and control groups. The index date for intervention group was defined as the most recent date of an opioid prescription prior to PGx testing. The index date for control group was defined as the earliest date of opioid prescription. All baseline covariates were collected in the one-year period prior to the index date. The outcome was a binary target variable, indicating whether the patient was in the intervention or control group based on their PGx test record. If a patient was in the intervention group, their outcome value was encoded as 1 and otherwise, 0.

### 2.2. Feature selection and data pre-processing

We selected four different categorizes of features for our study sample: (i) demographics, (ii) clinical history, (iii) medication use, and (iv) SDoH. Demographics included age, gender, and race, where age was continuous, sex was binary, and race was multi-category which was one-hot-encoded to get separate binary inputs for each racial group and prevent the ordinality assumption by the model. We used the international classification of disease (ICD-9 and ICD-10) codes to determine the clinical history variables, including a diagnosis of AIDS/HIV, alcohol abuse, blood loss anemia, cardiac arrhythmias, chronic pulmonary disease, coagulopathy, systolic (congestive) heart failure, deficiency anemia, depression, diabetes with complications, diabetes without complications, drug abuse, fluid and electrolyte disorder, hypertension, hypothyroidism, liver disease, lymphoma, metastatic cancer, neurodegenerative disorders, obesity, paralysis, peptic ulcer disease, peripheral vascular disease, psychosis, pulmonary circulation disorders, renal failure, rheumatoid arthritis/collagen, solid tumor without metastasis, valvular disease, and weight loss. Furthermore, we included Elixhauser comorbidity index [12] to calculate the overall healthcare burden for each patient. In addition, the SDoH information included average household size, Gini index as a measure of income inequality, median household income, and median rent, all linked to UF Health EHR based on year and ZIP code using the Agency for Healthcare Research and Quality (AHRQ) SDoH database [13]. We removed all the missing data, so all patients had complete information for model development.

### 2.3. Model development and hyperparameter optimization

We adopted a comprehensive strategy for model development using LR, XGBoost [14], RF [15], support vector machines (SVM) [16] with linear and radial basis function (RBF) kernels (i.e., LSVM and SVM-RBF), and NNs to capture predictive performance across a wide range of models for PGx uptake prediction. LR was first developed due to its ability to capture linear relationships in the data and providing baseline performance. Similarly, Naïve bayes (NB) model was developed due to its simplicity and effectiveness in high-dimensional spaces. However, considering LR’s and NB’s limitations in capturing complex non-linear relationships, we trained tree-based models, including XGBoost and RF, given their ability to capture non-linear interactions that may be critical to accurately predicting the PGx uptake. Linear SVM and SVM-RBF were developed to capture high-dimensional complex interactions among input features. Finally, NNs were trained to explore more complex interactions among features that may be missed by traditional models.

To train the models, we randomly partitioned the data with stratified 80/20 train/test split. To mitigate the risk of overfitting and get the best performance out of ML models, we conducted 5-fold cross validation. For LR, we tuned the parameter C with values of 0.01, 0.1, 1, and 10. To tune NB, we used smoothing parameters ranging from 10^−9^ to 10^−5^ with 10x increments. Both XGBoost and RF models were tuned with 100, 200, 300, 400, and 500 trees to explore the effect of different numbers of trees on performance. Linear SVM and SVM-RBF were optimized with C values equal to 0.01, 0.1, 1, and 10. Finally, NNs were trained with all possible combinations of various optimizers and learning rates. The optimizers included stochastic gradient descent (SGD) [17], Adam [18], and Nadam [19], and learning rates ranged from 10^−5^ to 10^−1^ with 10x increases. The best hyperparameters were selected based on the highest C-statistics achieved on the training set during cross-validation process.

We trained all the models using the best-performing hyperparameters and evaluated their performance on the test set. Notably, we employed a balanced class weighting approach, assigning proportionally higher weights to minority class, to decrease the risk of overfitting when training each of the models. To improve the performance and enhance generalizability of the models, we created a weighted probability-based ensemble classifier. To this end, we searched for the best individual model weights using AUC-ROC as the performance metrics, conditioned on having a total sum of weights equal to 1. This ensemble model was selected as the final best model for PGx uptake prediction.

### 2.4. Comprehensive evaluation

We followed TRIPOD+AI guidelines [20] to evaluate the performance of ML models. Area under receiver operating characteristics curve (AUC-ROC) or C-statistics demonstrates the discriminative ability of the model, considering the trade-off between sensitivity and specificity at different threshold values. DeLong test [21, 22] is used to identify whether there is a statistically significant difference between AUC values. Accuracy provides the overall performance of the model by calculating proportion of true negative and true positive cases compared to all the samples in the data. However, this measure may be misleading for rare events and not suitable for highly imbalanced datasets. To address this, we calculated specificity, which measures the model’s ability to correctly identify true negative cases and recall or sensitivity, indicating model’s performance for identifying true positive cases. Since model performance may not be optimal, we use Youden index [23–25] to obtain the best performing classification threshold for each model, where the highest value of ‘specificity+sensitivity-1’ is achieved. To further evaluate model performance for real-world applications, we calculated the number needed to evaluate and predictive positives per 100 patients, showing how many predictions must be made to identify one actual positive case and the number of PGx uptake predictions per 100 individuals, respectively. Moreover, we did stratification analysis, where PGx uptake probabilities were categorized into 10 deciles in ascending order and the percentage of actual PGx uptake in the data was identified in each decile to evaluate the ability of the model in identifying more uptakes in the higher strata. In addition, we used Shapley Additive exPlanations (SHAP) [26] analysis to report feature importance and identify the features that are most influential to PGx uptake.

## 3. Results

We used data from UF Health IDR with patients from a wide of range of demographic and clinical backgrounds. The data included a total of 455,773 patients, 7,645 (1.68%) with a recorded PGx test and 448,128 (98.32%) without. After removing 160,914 further patients who had no opioid prescription, there were 294,859 patients using opioids in the cohort. We further excluded patients with no race information and SDoH variables (after matching with AHRQ database). Overall, the final cohort included 242,640 patients, where 3,510 had a recorded PGx uptake and 239,130 did not. The cohort had an average age of 58 with a standard deviation of 17.47 years, a gender distribution comprising 41.49% as males and 58.51% as females, and a racial distribution of 64.94% White, 28.77% Black, and 6.29% belonging to other racial groups (i.e., Hispanic, White Hispanic, Black Hispanic, Asian, Pacific Islander, American Indian, Multiracial, and other). In addition, the cohort had an average household size of 2.58, a Gini index of 0.45, and an average median income of $46,324.

Table 1 presents the key sociodemographic characteristics of patients in the cohort. Moreover, the distribution of patient characteristics in the training and test sets are provided in Table 2.

**Table 1.**
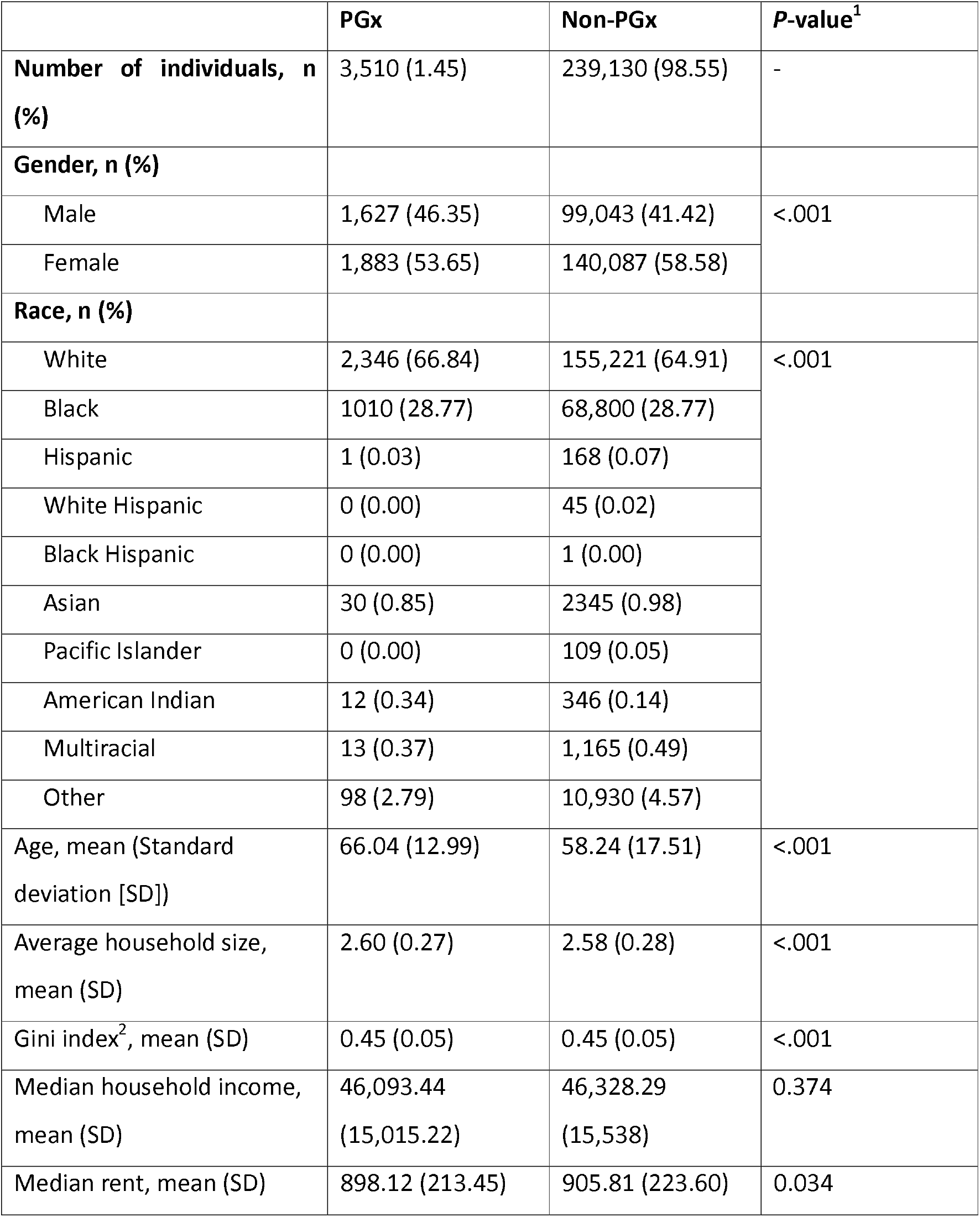
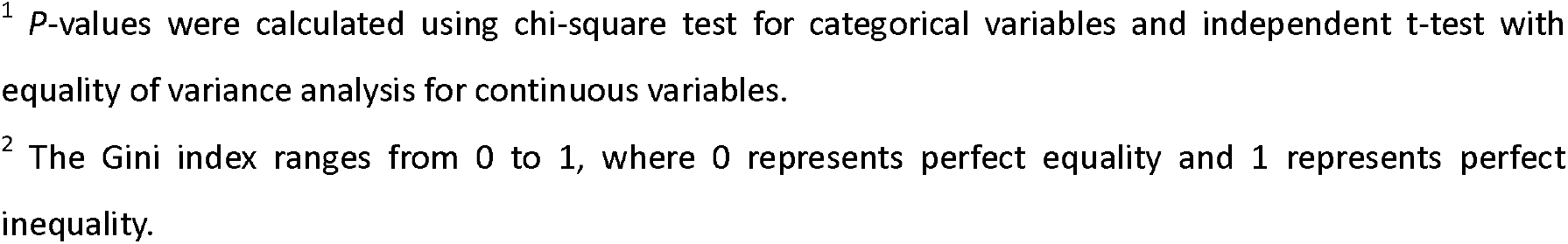
Sociodemographic summary of patients.

|  | PGx | Non-PGx | P-value<sup>1</sup> |
| --- | --- | --- | --- |
| **Number of individuals, n (%)** | 3,510 (1.45) | 239,130 (98.55) | - |
| **Gender, n (%)** |  |  |  |
| Male | 1,627 (46.35) | 99,043 (41.42) | <.001 |
| Female | 1,883 (53.65) | 140,087 (58.58) |  |
| **Race, n (%)** |  |  |  |
| White | 2,346 (66.84) | 155,221 (64.91) | <.001 |
| Black | 1010 (28.77) | 68,800 (28.77) |  |
| Hispanic | 1 (0.03) | 168 (0.07) |  |
| White Hispanic | 0 (0.00) | 45 (0.02) |  |
| Black Hispanic | 0 (0.00) | 1 (0.00) |  |
| Asian | 30 (0.85) | 2345 (0.98) |  |
| Pacific Islander | 0 (0.00) | 109 (0.05) |  |
| American Indian | 12 (0.34) | 346 (0.14) |  |
| Multiracial | 13 (0.37) | 1,165 (0.49) |  |
| Other | 98 (2.79) | 10,930 (4.57) |  |
| Age, mean (Standard deviation [SD]) | 66.04 (12.99) | 58.24 (17.51) | <.001 |
| Average household size, mean (SD) | 2.60 (0.27) | 2.58 (0.28) | <.001 |
| Gini index<sup>2</sup>, mean (SD) | 0.45 (0.05) | 0.45 (0.05) | <.001 |
| Median household income, mean (SD) | 46,093.44<br>(15,015.22) | 46,328.29<br>(15,538) | 0.374 |
| Median rent, mean (SD) | 898.12 (213.45) | 905.81 (223.60) | 0.034 |

<sup>1</sup> *P*-values were calculated using chi-square test for categorical variables and independent t-test with equality of variance analysis for continuous variables.
<sup>2</sup> The Gini index ranges from 0 to 1, where 0 represents perfect equality and 1 represents perfect inequality.

**Table 2.** Distribution of patient characteristics in the training and test sets.

| Characteristics | Training<br>(n=194,112) | Testing<br>(n=48,528) |
| --- | --- | --- |
| **Gender, n (%)** |  |  |
| Male | 80,519 (41.48) | 20,151 (41.52) |
| Female | 113,593 (58.52) | 28,377 (58.48) |
| **Race, n (%)** |  |  |
| White | 125,938 (64.88) | 31,629 (65.18) |
| Black | 55,993 (28.85) | 13,817 (28.47) |
| Hispanic | 128 (0.07) | 41 (0.08) |
| White Hispanic | 33 (0.02) | 12 (0.02) |
| Black Hispanic | 1 (0.00) | 0 (0.00) |
| Asian | 1,877 (0.97) | 498 (1.03) |
| Pacific Islander | 89 (0.05) | 20 (0.04) |
| American Indian | 273 (0.14) | 85 (0.18) |
| Multiracial | 919 (0.47) | 259 (0.53) |
| Other | 8,861 (4.56) | 2,167 (4.47) |
| Age, mean (Standard deviation [SD]) | 58.36 (17.48) | 58.32 (17.45) |
| Average household size, mean (SD) | 2.58 (0.28) | 2.58 (0.28) |
| Gini index, mean (SD) | 0.45 (0.05) | 0.45 (0.05) |
| Median household income, mean (SD) | 46,327.05<br>(15,529.07) | 46,316.28<br>(15,536.70) |
| Median rent, mean (SD) | 905.64 (223.38) | 905.92 (223.78) |

### 3.1. Model performance

The ROC curves of all models and their corresponding AUC or C-statistics with 95% confidence intervals (CIs) are provided in Fig. 2. The ensemble model achieved the highest C-statistics of 79.61% using 0.7 and 0.3 weights for XGB and SVM-RBF, respectively. In contrast, NB achieved the lowest C-statistics at 72.49%. Other models had comparable C-statistics, equal to 75.38% (RF), 76.72% (LR), 76.46% (LSVM), 77.73% (SVM-RBF), 78.05% (NN), and 78.94% (XGB). Additionally, the DeLong test results indicated that there was a statistically significant difference (p <0.05) between the C-statistics of ensemble model and all other classifiers, showing its higher performance is less unlikely to be due to chance.

**Fig. 2.**
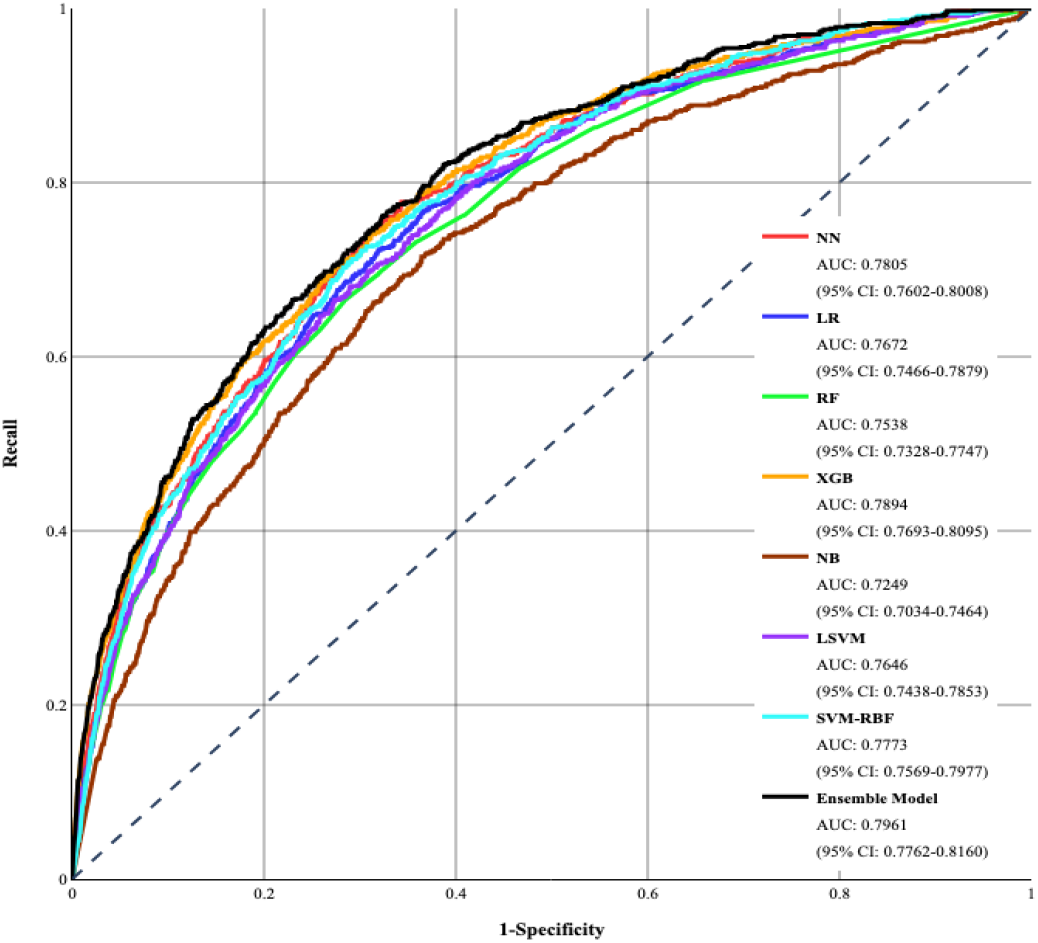
ROC curves of developed models.

Since model performance was not optimal, we used Youden index [23–25] to obtain the best performing classification threshold for each model. Fig. 3 portrays the confusion matrices for all developed models based on their Youden index threshold. Table 3 provides a summary of performance for all developed models. Accuracy representing the proportion of correctly classified cases, was highest for XBG (72.54%), followed by RF (71.31%), LSVM (71.10%), and SVM-RBF (70.60%). Other classifiers had 65-70% accuracy with ensemble model having an accuracy of 67.38%.

**Table 3.**
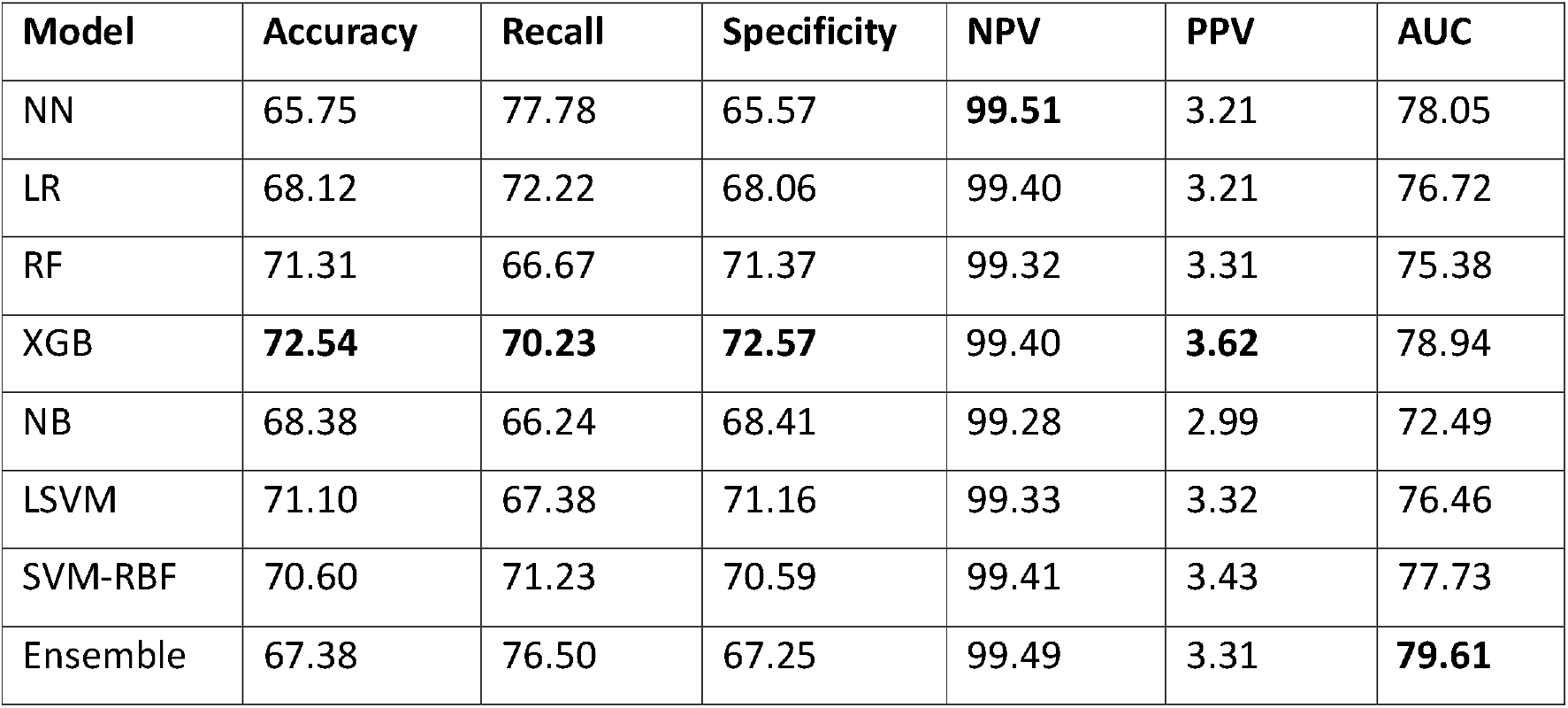
Pefromance metrics of developed models based on Youden index.

| Model | Accuracy | Recall | Specificity | NPV | PPV | AUC |
| --- | --- | --- | --- | --- | --- | --- |
| NN | 65.75 | 77.78 | 65.57 | **99.51** | 3.21 | 78.05 |
| LR | 68.12 | 72.22 | 68.06 | 99.40 | 3.21 | 76.72 |
| RF | 71.31 | 66.67 | 71.37 | 99.32 | 3.31 | 75.38 |
| XGB | **72.54** | **70.23** | **72.57** | 99.40 | **3.62** | 78.94 |
| NB | 68.38 | 66.24 | 68.41 | 99.28 | 2.99 | 72.49 |
| LSVM | 71.10 | 67.38 | 71.16 | 99.33 | 3.32 | 76.46 |
| SVM-RBF | 70.60 | 71.23 | 70.59 | 99.41 | 3.43 | 77.73 |
| Ensemble | 67.38 | 76.50 | 67.25 | 99.49 | 3.31 | **79.61** |

**Fig. 3.**
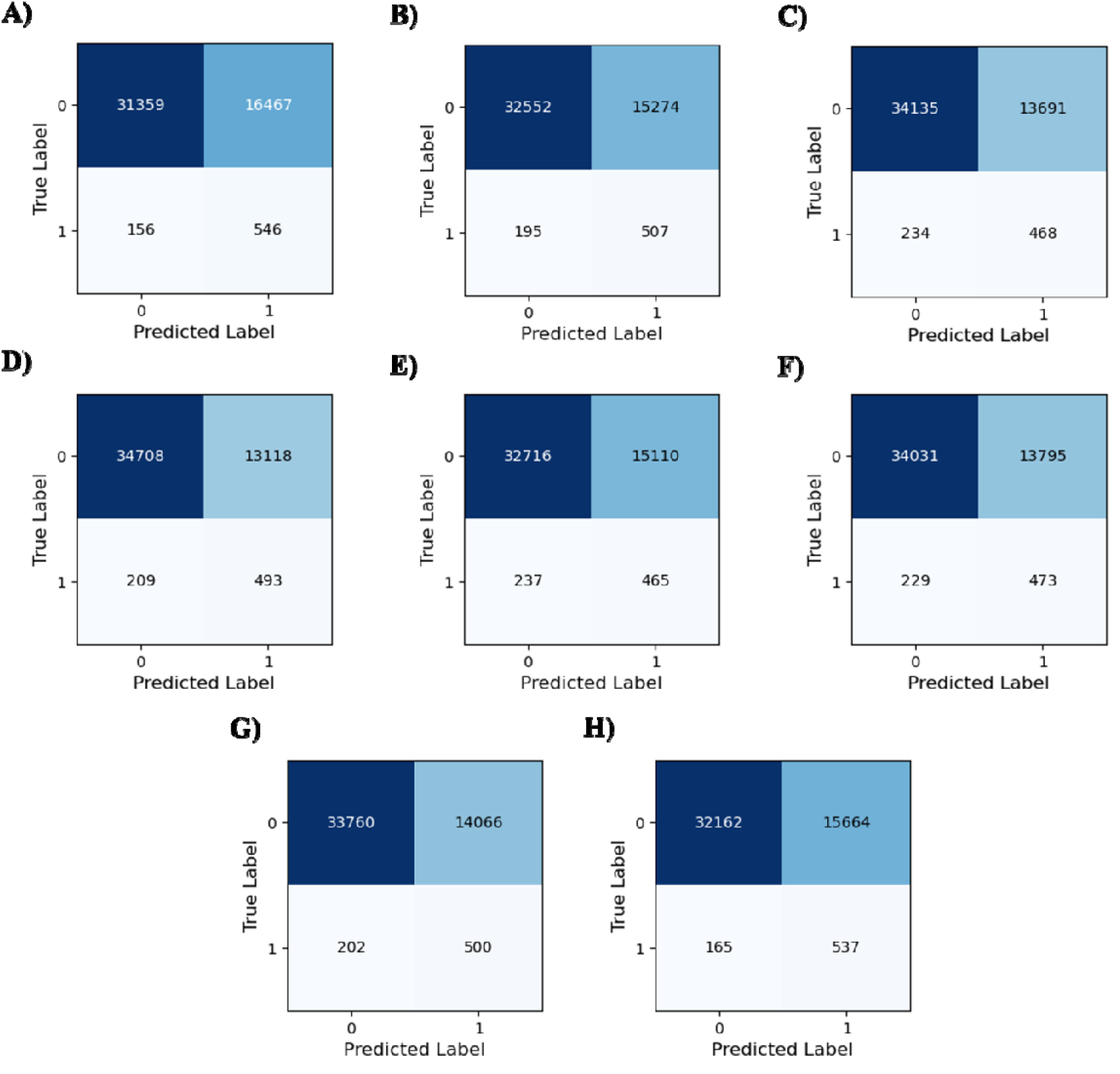
Confusion matrix of models based on Youden index. (A) NN; (B) LR; (C) RF; (D) XGB; (E) NB; (F) LSVM; (G) SVM_RBF; (H) Ensemble model.

To assess the model performance in identifying true positive cases (i.e., those with a recorded PGx uptake) and true negative cases (i.e., those without a recorded PGx uptake), recall and specificity were calculated. Recall values ranged from 66.24-77.78% with NN model achieving the highest recall and ensemble model achieving 76.50%. Specificity values ranged from 65.57-72.57%, with XGB model achieving the highest specificity. To further assess models’ performance in distinguishing between true positives and true negatives, negative predictive value (NPV) and positive predictive value (PPV) were calculated. NPV values ranged from 99.28-99.51% with NN model having the highest NPV, while PPV values ranged from 2.99-3.43% with SVM-RBF achieving the highest value.

Stratification analysis results are presented in Fig. 4. The analysis was performed by dividing the predicted probabilities of PGx uptake into deciles and calculating observed uptake rates within each to assess performance across the probability groups. The lowest decile was decile 1 with lowest probability values, while decile 10 represented the highest probability values. Within each decile, observed uptake rates were calculated to plot against the probability deciles. The observed event rates demonstrated an increasing trend across the probability deciles (except for the 5^th^). The event rate in the highest probability decile was 6.59% while 0.12% in the lowest decile.

**Fig. 4.**
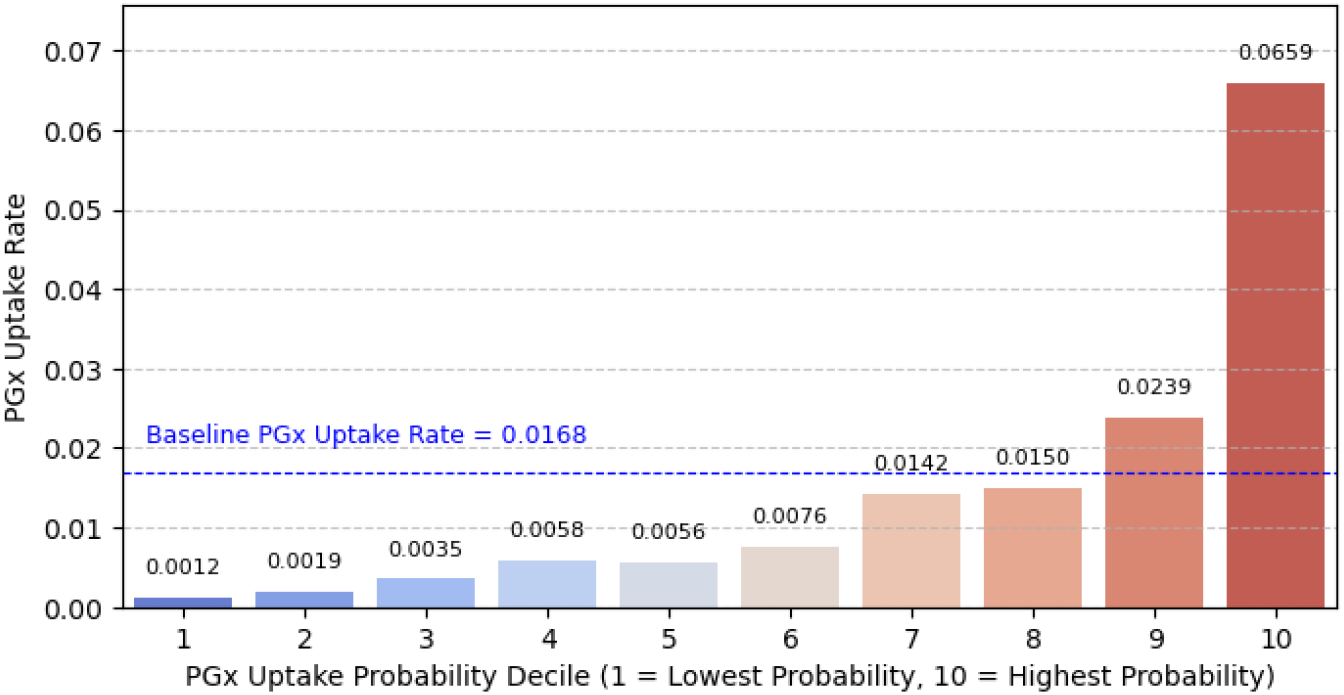
Stratification analysis for the probability of PGx uptake.

Fig. 5 demonstrates the plots of recall values against predictive positives per 100 and number needed to evaluate. Predictive positives per 100 represents the number of individuals per 100 who are predicted to undergo PGx across different recall values. The number needed to evaluate indicates the number of individuals that need to be assessed by the model in order to find one true positive (i.e., recorded PGx uptake).

**Fig. 5.**
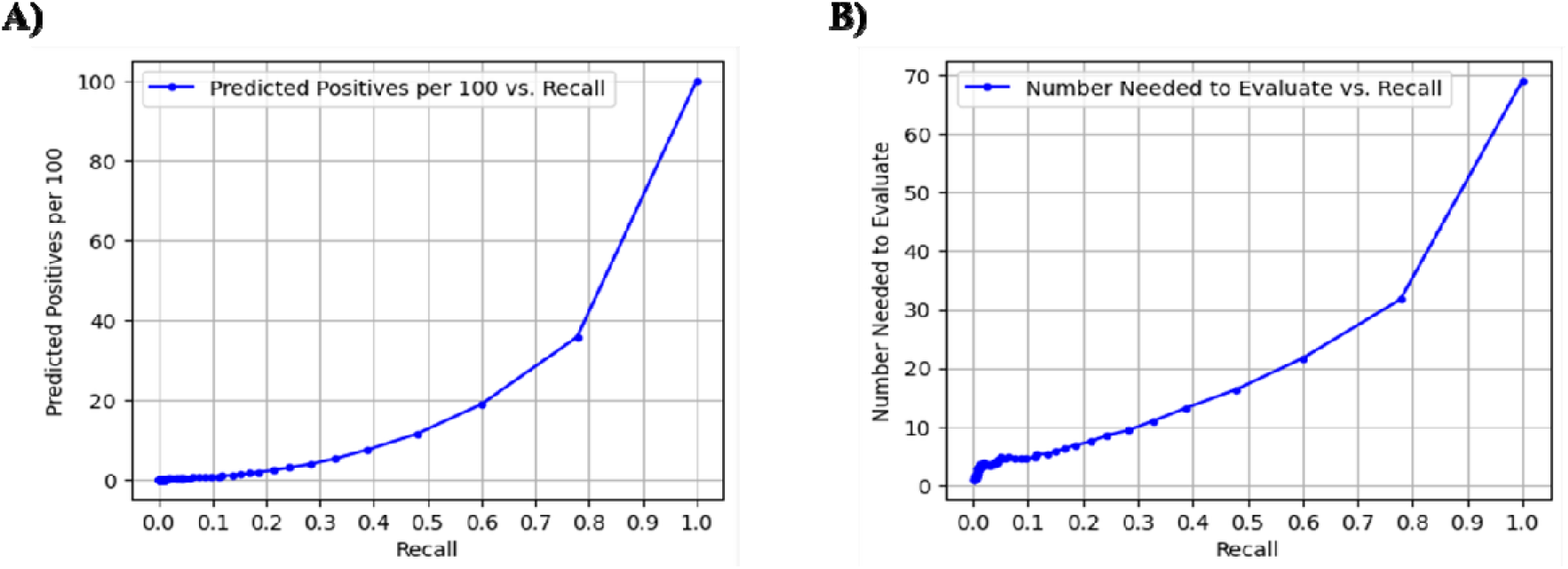
Recall vs. (A) predicted positives per 100, (B) number needed to evaluate.

SHAP analysis for feature importance using the XGB model has been shown in Fig. 6. Age had the highest impact on PGx uptake, where older patients are more likely to undergo PGx. Hypertension and median household income were the next most important features, both associated with a higher likelihood of PGx uptake. Other SDoH factors were among the top 7 important factors for PGx uptake. In contrast, racial group variables (i.e., Hispanic, Pacific Islander, and White Hispanic) had the lowest influence on the outcome.

**Fig. 6.**
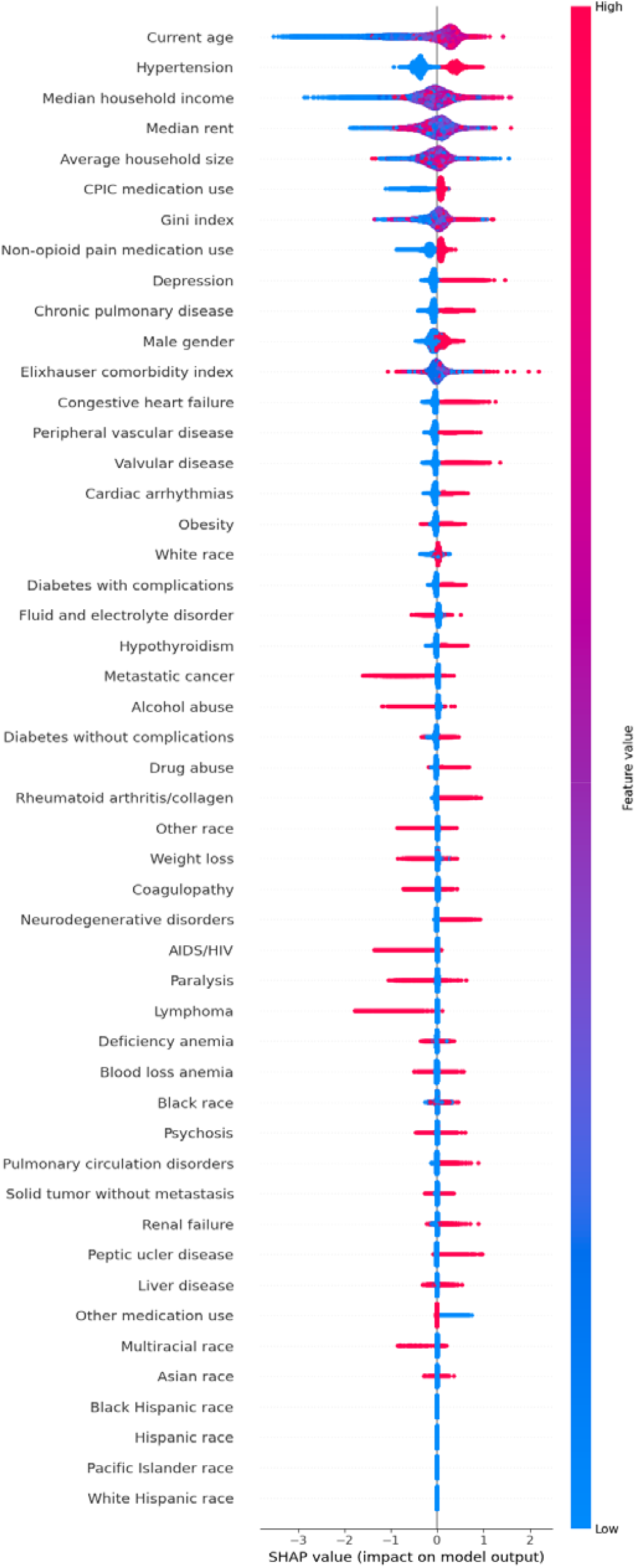
SHAP analysis for feature importance (The x-axis shows the impact on prediction and the color represents the feature value (i.e., red for high and blue for low)).

## Discussion

The results of this study demonstrate that the ensemble model achieved the highest AUC compared to other models. Although the XGB model, when evaluated at its Youden index performed better than other models in terms of accuracy, recall, specificity, and NPV, we selected the ensemble model as the final classifier. This was because the ensemble model harnesses the strengths of both high-performing XGB and SVM-RBF models for prediction. It is notable that although the study used PPV as a performance metric, it is highly dependent on outcome prevalence and may not generalize across different patient populations or clinical settings, reducing its applicability in decision-making. Overall, the ensemble model proves to be a viable tool for accurately predicting PGx uptake, thereby supporting more informed, data-driven decision-making in clinical settings. By providing precise and individualized PGx uptake probabilities, clinicians will be able to identify individuals who are less likely to undergo PGx testing. By prioritizing these patients, the model has the potential to assist clinicians in optimizing resource allocation based on patient needs and facilitating PGx uptake, ultimately improving patients’ pain management outcomes.

The dataset used for model development exhibited class imbalance, with a relatively low percentage of patients having recorded PGx uptake. Despite utilizing 5-fold CV for hyperparameter optimization and a balanced weighting approach for final model development, this imbalance resulted in a lower PPV compared to other performance metrics. This means the model has a higher number of false positives, meaning that many of patients who are predicted to undergo PGx testing will not, potentially leading to inappropriate opioid treatment for the patients. Conversely, the class imbalance led to a very high NPV, meaning that most no-uptake predictions are accurate. This high NPV could be advantageous in clinical decision-making, as it allows for the allocation of more resources and encouragement plans for patients predicted not to undergo PGx testing. By identifying characteristics of these patients, targeted strategies can be developed to increase their PGx uptake rate, ultimately leading to improved patient outcomes.

This study utilized various clinical and demographic variables to develop more-informed ML models. However, the underrepresentation of certain racial groups, such as Hispanic and Pacific Islander could potentially affect models’ outcomes. This raises concerns about the bias and fairness issues, as ML models may exhibit lower performance for these groups compared to others. Additionally, while the study incorporated SDoH information as potential predictors of PGx uptake, this information was based on ZIP code-level data, which may lack the necessary granularity required for precise prediction. Detailed individual-level data on variables such as income, education, and household size could enhance model’s reliability and improve the accurate identification of each feature’s contribution on the outcome.

Another limitation of the study is its generalizability to other populations in different clinical settings. The model’s performance may not directly translate to external populations with varying demographic and clinical backgrounds. Specifically, the model was trained on data from a single EHR system including minority racial groups and the results may not be applicable to other racial groups. Therefore, caution is warranted when interpreting the model’s outcomes for these populations. In addition, some of the patients in the data who had a recorded PGx uptake were part of a clinical trial where the cost of testing was covered. This would potentially influence the ability of the model in identifying the true impact of patients’ economic and income status on undergoing PGX testing. To demonstrate model’s broader applicability, future work should focus on external validation using independent datasets from different clinical settings. This would help evaluate model’s robustness and clinical utility for real-world decision-making.

The SHAP value analysis using the XGB model highlighted the contribution of each feature to individual predictions, enhancing model’s clinical credibility and trustworthiness. The most influential features were age, hypertension, and household income, suggesting that these features require more attention for increasing PGx uptake among patients. Notably, PGx uptake is not necessarily due to opioid prescribing and could be due to prescribing of other medications and participation in a research project, potentially affecting the study results. Thus, although the results demonstrate model’s transparency and clinical utility, more research is needed before integrating the model into a real-world clinical decision support system. While SHAP diagram ranks features based on their overall importance, it is crucial to examine critical factors for each patient on a case-by-case basis when making clinical decisions. Overall feature importance does not always reflect individual patient’s conditions and their influence on the outcome.

The uptake stratification analysis showed that the ensemble model effectively differentiated patients across PGx uptake deciles, with higher uptake probabilities corresponding to higher observed uptake rates. Such an analysis can assist clinicians in categorizing patients into different deciles based on their likelihood of PGx uptake and prioritize those in lower deciles (less likely for the PGx uptake) for optimal resource allocation and improved access to testing. In addition, it can help clinicians authorize insurance coverage and provide post-test consultation for patients in higher uptake deciles. While there was an overall increasing trend in uptake rates as the probabilities increased, there was a slight decrease (i.e., 0.002) in 5^th^ decile, indicating that the higher probabilities in this decile could not capture a higher uptake rate. In addition, stratification analysis often relies on pre-defined thresholds (i.e., 10% in this study) to categorize probabilities into deciles, which may not be aligned with the way clinicians make decisions and could limit its utility in clinical decision-making. Despite these limitations, the stratification analysis supports the potential use of the ensemble model in prioritizing patients based on their likelihood of undergoing PGx, leading to optimized resource allocation and improved patient outcomes.

The implementation of the PGx uptake prediction model addresses the barriers to PGx testing in opioid therapy. By providing probability of PGx uptake at the individual patient level, clinicians can strategically optimize resource allocation across their patient population. For patients with a high predicted probability of PGx uptake, the clinical workflows can be expedited by focusing on authorizing insurance coverage or allocating clinical pharmacist time for post-test consultation. This efficient deployment of resources where they are most likely to be utilized allows practices to simultaneously redirect access support toward patients with lower uptake probabilities. Such support may include patient education on the benefits and implications of PGx testing or addressing financial barriers. The model’s ability to stratify patient’s likelihood of PGx uptake transforms the current trial-and-error paradigm of opioid prescribing into a more systematic framework for clinical decision-making.

## Conclusions

PGx is a viable tool for matching patient’s genetic profile to suitable opioids for pain treatment. This study proposed ML models for PGx uptake prediction using data from an EHR system. Results demonstrated that the ensemble ML model combining XGB and SVM-RBF classifiers achieved the highest AUC at 79.61%, making it a reliable prediction model for PGx uptake prediction. Additionally, the uptake stratification and feature importance analysis using SHAP values further indicated model’s utility for real-world applications. Following further validation using external dataset, this model can be integrated into a clinical decision support system, enabling clinicians in more informed, risk-averse clinical decisions, ultimately improving patient outcomes.

## Data Availability

The data cannot be made available to the readers.

